# From Keywords to Context: Bridging Expert Insight and Language Models for Multidimensional Sleep Health Classification in Clinical Notes

**DOI:** 10.1101/2025.06.06.25329167

**Authors:** Syed-Amad Hussain, Ariana Calloway, Joseph W Sirrianni, Eric Fosler-Lussier, Mattina Davenport

**Affiliations:** Nationwide Children’s Hospital; The Ohio State University

## Abstract

Accurate detection of multidimensional sleep health (MSH) information from electronic health records (EHRs) is critical for improving clinical decision-making but remains challenging due to sparse documentation and class imbalance. This study investigates whether integrating expert-guided annotations and keyword-based heuristics with large language models (LLMs) enhances the extraction of nuanced MSH indicators from clinical narratives. Using a novel, expertly annotated dataset (NCH-Sleep), we trained and evaluated models to classify clinical notes across nine clinically relevant MSH categories. Our baseline model demonstrated substantial predictive capability using raw text alone. Incorporating manually annotated spans (oracle annotations) dramatically improved performance, highlighting the benefit of targeted expert guidance. Additionally, employing curated keyword annotations within varying context windows significantly enhanced model interpretability while retaining strong predictive accuracy. Through detailed bias analyses, we identified consistent performance across demographics and clinical settings, although specific disparities underscored the importance of balanced expert oversight. Our findings emphasize the value of expert-informed supervision and heuristic approaches in building scalable, interpretable clinical NLP systems for sleep health classification.

## Introduction

Large language models (LLMs) offer a promising avenue for extracting complex clinical phenomena from unstructured electronic health record (EHR) text at scale. One such area is multidimensional sleep health (MSH), a multifaceted concept encompassing not only sleep disorders but also behavioral, temporal, and quality-related aspects of sleep. Reliable identification of MSH-related information from clinical narratives has the potential to support the development of targeted detection tools, streamline patient cohort selection for research and interventions, and ultimately improve clinical decision-making. However, this task is complicated by substantial class imbalance, varying documentation practices, and the inherent sparsity of explicit mentions across MSH categories.

In this work, we tackle two critical challenges in MSH detection: first, achieving robust, high-accuracy classification of clinical notes into nine clinically relevant MSH categories, and second, enhancing the interpretability of model decisions so that outputs are both reliable and actionable for clinicians. Our objectives are to develop a high-performing classifier that can extract nuanced MSH signals from free text without relying on extensive external context, and to rigorously evaluate the impact of expert knowledge on model performance and interpretability. To this end, we compare model-derived features with expert-provided signals, including manually annotated spans (oracle annotations) and curated keyword lists, to examine how these forms of supervision alter predictions and generalizability.

We begin by establishing a baseline classifier that uses only the unmodified clinical note along with a brief natural language description of the target class. Building on this baseline, we then incorporate expert knowledge in two forms: (i) oracle annotations, which consist of manually highlighted spans that justify MSH classifications, and (ii) curated keyword lists that can either narrow the input context to clinically relevant segments (using both large and small context windows) or simply highlight key terms within the note. These experiments allow us to evaluate not only predictive performance but also the degree of overlap between expert-derived signals and patterns identified by the model. Importantly, by comparing different context window sizes for the keyword-based guidance, from expansive windows that optimize predictive performance to smaller, more digestible windows that enhance interpretability, we assess the trade-off between raw predictive accuracy and the clarity of model outputs.

Ultimately, this study provides insights into how expert supervision and domain-specific heuristics can be integrated with LLM-based classifiers to optimize MSH extraction. The findings have significant implications for clinical use cases: for scenarios demanding the highest predictive performance, broad context inclusion via large windows may be most effective, whereas in settings designed for human-in-the-loop decision-making, concise keyword annotations (e.g., using small windows) can yield comparable accuracy while enhancing interpretability. This balance between automation and clinician engagement is critical for ensuring that decision support systems are not only effective but also trusted and readily adopted in real-world clinical workflows.

## Background

Natural language processing (NLP), particularly through large language models (LLMs), has emerged as an effective method for extracting complex health information from clinical narratives. Prior approaches to clinical NLP commonly relied on rule-based algorithms and manually selected keywords. For instance, Horner et al. (2022) utilized a keyword-driven approach to detect sleep-related complaints in primary care notes, which achieved high specificity but limited sensitivity, mainly due to reliance on fixed keywords and simple pattern-matching strategies. Similarly, Sivarajkumar et al. (2024) showed superior performance of keyword-based methods compared to traditional machine learning models when extracting various sleep-related mentions from clinical text, achieving F1-scores ranging from 0.91 to 1.0. Extending these approaches, Sirrianni et al. (2025) refined sleep vocabularies using ontologies and semantic embeddings, significantly improving recall for identifying notes with sleep mentions; however, this approach did not specify exact textual locations or distinguish among sleep-related subcategories.

Recent developments in pretrained language models have demonstrated improvements in extracting clinical information by capturing context and semantics more effectively than keyword-based methods alone. For example, Li et al. (2024) utilized domain-adapted (e.g., BioClinicalBERT) and task-specific models (e.g., MentalBERT) to identify suicide-related behaviors from psychiatric notes, demonstrating clear advantages over generic models. This work underscored the potential of models pretrained on in-domain clinical or mental health-specific data to improve accuracy on specialized tasks. Moreover, Li et al. (2024) adopted a binary classification approach, converting complex multi-label problems into simpler, independent binary tasks, which improved model interpretability and performance.

In the specific domain of multidimensional sleep health (MSH), Hussain et al. (2025) empirically evaluated various LLM strategies for classifying free-text clinical notes into categories such as sleep behaviors, quality, timing, medications, and disorders. Their results indicated that fine-tuned discriminative models consistently outperformed generative models relying solely on prompting, emphasizing that explicit fine-tuning provides improved accuracy and computational efficiency. Additionally, Hussain et al. (2025) showed that classification performance can substantially benefit from increased quantities of domain-specific training data, suggesting scalability improvements through dataset expansion.

Collectively, these studies illustrate the ongoing transition from keyword-based NLP methods toward context-aware, fine-tuned LLMs for clinical information extraction tasks. While keyword strategies provide a straightforward, interpretable baseline, advanced LLM-based methods can capture subtle and context-dependent clinical phenomena more effectively.

However, leveraging expert knowledge to guide LLMs, particularly by integrating domain-informed keywords or expert annotations, remains an open question for maximizing model accuracy, interpretability, and clinical usability.

## Data

Our work primarily employs an internal sleep mentions dataset. All datasets originate from electronic medical records, and each note within these datasets has been annotated to indicate the presence or absence of specific classes.

### NCH-Sleep Dataset

We developed a novel sleep-focused dataset, NCH-Sleep, comprising 1200 clinical documents authored between 2018 and 2023 at Nationwide Children’s Hospital (NCH). The initial pool of clinical records was retrieved from the hospital’s electronic health records (EHR) system through keyword searches guided by an expert-curated list of sleep-related terms. These records spanned four distinct departments: primary care, behavioral health, healthy weight, and school-based health programs. From approximately 700,000 available notes, we selected a random yet stratified sample of 1200 documents, ensuring balanced representation across departments and years. Each selected note underwent expert annotation to determine the presence of nine distinct sleep-related Medical Subject Headings (MSH) categories, with annotators marking relevant textual evidence. Detailed descriptions of these nine sleep categories are provided in Table 2, while Table 3 summarizes the frequency distribution of clinical notes across each department.

**Table 1:**
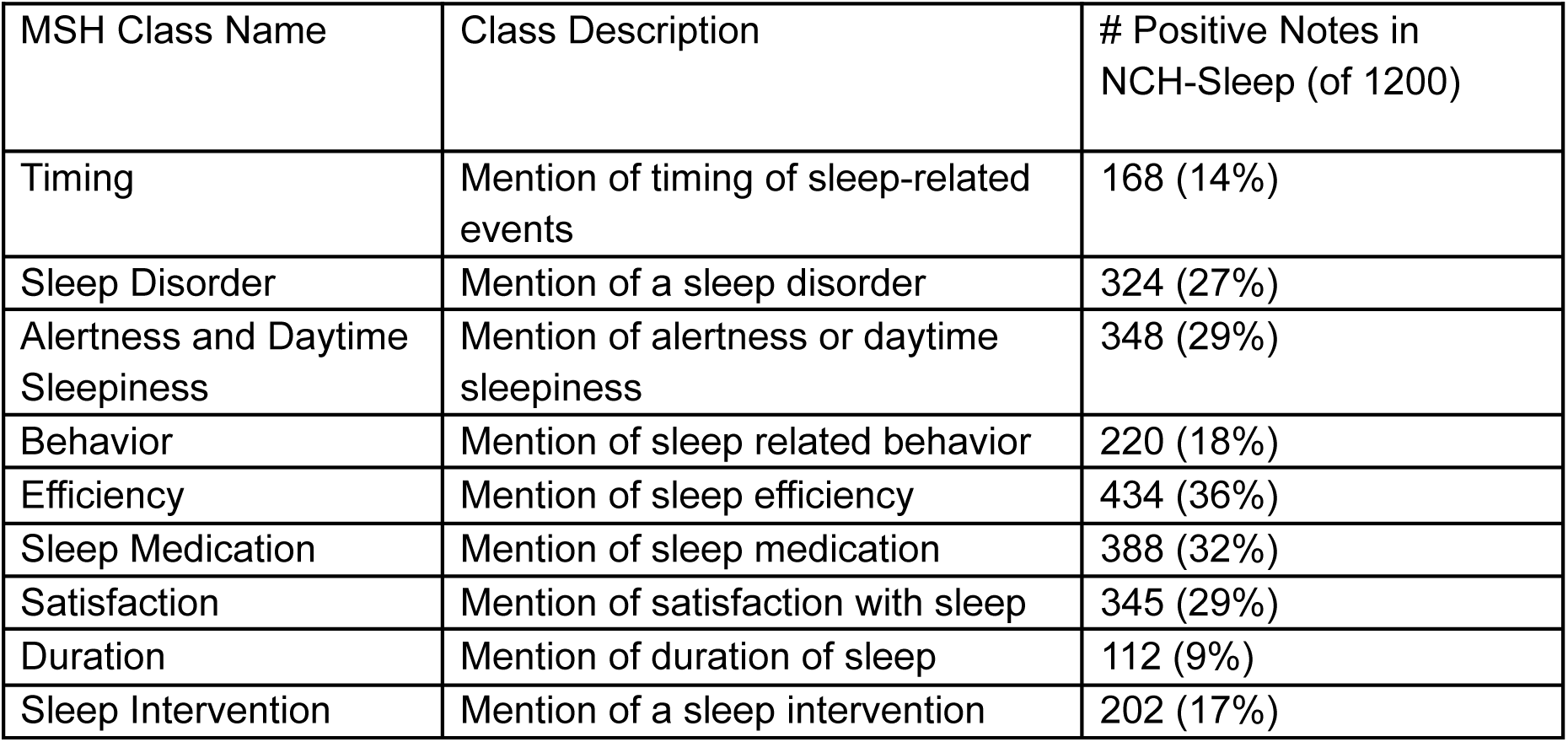
The 9 MSH classes annotated in the NCH-Sleep dataset and evaluated in each of our experiments. Class descriptions are used to help inform models.

| MSH Class Name | Class Description | # Positive Notes in NCH-Sleep (of 1200) |
| --- | --- | --- |
| Timing | Mention of timing of sleep-related events | 168 (14%) |
| Sleep Disorder | Mention of a sleep disorder | 324 (27%) |
| Alertness and Daytime Sleepiness | Mention of alertness or daytime sleepiness | 348 (29%) |
| Behavior | Mention of sleep related behavior | 220 (18%) |
| Efficiency | Mention of sleep efficiency | 434 (36%) |
| Sleep Medication | Mention of sleep medication | 388 (32%) |
| Satisfaction | Mention of satisfaction with sleep | 345 (29%) |
| Duration | Mention of duration of sleep | 112 (9%) |
| Sleep Intervention | Mention of a sleep intervention | 202 (17%) |

**Table 2:** Departments and their respective number of notes within NCH-Sleep.

|  |  |
| --- | --- |
| Department | # Notes in NCH-Sleep |
| Primary Care | 691 |
| Behavioral Health | 428 |
| Healthy Weight | 57 |
| School Based | 24 |

**Table 3:** Results for our baseline models trained on NCH-Sleep-Train and evaluated on NCH-Sleep-Test. Findings from Hussain et al. (2025) are reported for comparison.

| Model | Precision | Recall | F1 | AUROC |
| --- | --- | --- | --- | --- |
| Hussain et al. (2025) | 0.48 | 0.19 | 0.27 | 0.65 |
| ModernBERT-large | 0.35 | 0.58 | 0.44 | 0.72 |
| Gemma-2-2b | 0.53 | 0.77 | 0.62 | 0.86 |

For all experiments reported here, the NCH-Sleep dataset was partitioned into training and testing subsets, comprising 900 and 300 notes, respectively (75% training, 25% testing). To enhance reliability, the 300-note test subset was independently annotated by two expert reviewers, with any disagreements resolved through a subsequent adjudication process. In contrast, each note in the 900-note training set received annotation from a single expert reviewer following the establishment of annotation guidelines and standards derived from the adjudication step. Although annotations in the training subset might exhibit less consistency due to the single-reviewer approach, this process simulates practical scenarios where extensive multi-reviewer annotation might not be feasible during model development.

The average note length is 5697 characters (4090 std dev) and an average of 2.7 positive MSH classes (2.0 std dev) per note.

### Task Structure

#### Boolean Classification by Class

Our evaluation, though conceptually similar to a multi-class classification scenario involving nine MSH classes, treats each class as conditionally independent due to the possibility of multiple simultaneous class labels for a single clinical note. This conditional independence simplifies the task by turning it into individual binary classification problems (true/false) for each class. Such framing allows models to focus specifically on predicting one class at a time, thereby reducing potential confusion between overlapping classes. Moreover, this structure facilitates querying models with classes that were not part of the original training set, providing clinicians flexibility in adapting the evaluation to new or evolving clinical needs and assessing model generalization capabilities. We present an evaluation of binary versus multiclass performance in Appendix A.1. Each binary classification example adheres to the following format:

~~~
Input: {‘clinical_note’: {*context_string}, ‘class_label’: {class_name}, ‘class_description’: {class_description}}
Output: 1/0
~~~

The variable *context_string is adjusted between our experiments, while all other variables are identical throughout the experiments in this work. In each experiment, we jsonify the input dictionary to get a string, which we provide to each model as input.

#### Negative Sampling Strategy for Training

Each note in our datasets is annotated explicitly with its positive class labels. Any class present in the dataset but not annotated as positive for a particular note is treated as a negative example. Introducing negative samples alongside positive examples creates a contrastive training environment, which guides models to differentiate more clearly between positive and negative class distributions, enhancing classification performance.

Our negative sampling reflects the expected clinical use case in that we query each note for all 9 MSH classes. Since notes on average contain 2.7 positive classes, this leaves an average of 6.3 negative classes per note, or an average ratio of 2.3 negative examples per positive example throughout our NCH-Sleep dataset.

## Experiment 1: Establishing a Baseline Classifier

To assess the extent to which LLMs can independently extract MSH information from clinical text, we first establish baseline performance using a set of fine-tuned models. This experiment evaluates the models’ ability to classify clinical notes into the nine predefined MSH categories using only two inputs: the raw text of the clinical note and a brief natural language description of the target class. No additional supervision, such as expert-labeled spans or curated keyword guidance, is introduced at this stage.

The goal of this phase is twofold. First, it allows us to quantify how much signal relevant to MSH can be learned directly from unstructured EHR narratives through standard fine-tuning. Second, it provides a benchmark against which later experiments incorporating expert knowledge can be compared. Improvements in classification performance here are attributed primarily to the model’s capacity to extract implicit patterns in the data without relying on external context. This establishes a starting point for understanding the value added by more structured forms of guidance in subsequent analyses.

The best-performing model from this experiment serves as the foundation for downstream experiments. By comparing this baseline to models that integrate expert-derived annotations or keyword heuristics, we aim to isolate the contribution of expert knowledge and explore the alignment between model-learned features and clinically meaningful indicators of sleep health.

### Methods

All models are trained on the 900-note train split of NCH-Sleep. Each note has 9 binary classification examples, one for each MSH class, as according to the task definition. In this experiment, we provide the whole, unmodified clinical note as the *context_string.

~~~
Input: {‘clinical_note’: {*whole_clinical_note}, ‘class_label’: {class_name}, ‘class_description’: {class_description}}
~~~

Each model is validated and tested on the 300-note, double expert adjudicated, test split of NCH-Sleep. We report precision, recall, and F1 score based on fine-tuned model predictions over NCH-Sleep-Test given a probability threshold of 0.5 for the positive class. We additionally report the ROC curve and AUROC, calculated using the raw positive-class probabilities provided by the fine-tuned models.

#### Training Environment

All models are trained on an NVIDIA H100 GPU with 80 GB of VRAM. We optimize Binary Cross Entropy (BCE) loss using the AdamW optimizer and a dropout rate of

0.3 applied to the final prediction layer. To account for class imbalance between MSH-present and MSH-absent cases, we apply a positive class weight of 3 in the BCE loss calculation. Each model is trained for up to 50 epochs, with early stopping triggered after 15 consecutive epochs without improvement on the validation loss. We select the epoch with the best validation loss for evaluation.

### Models

We evaluate two LLMs with distinct design tradeoffs: ModernBERT-large, a 395-million parameter encoder-only model, and Gemma-2-2b, a 2-billion parameter decoder-only model. These models were chosen due to their high performance on prior clinical classification tasks (Pandit et al. 2025). By comparing these models, we aim to identify whether the task benefits more from model scale and rich pretraining or from leaner architectures that are easier to adapt in resource-constrained settings.

#### ModernBERT-large

For this experiment, we fine-tune answerdotai/ModernBERT-base, a bidirectional encoder model that extends the original BERT architecture through modern optimizations and pretraining on over 2 trillion tokens (Warner et al. 2024). It is designed for efficiency, offering both fast runtime and a maximum sequence length of 8192 tokens, making it well-suited for long-form clinical text.

Previous work on MSH classification (Hussain et al. 2025) demonstrated that, with adequate training data, ModernBERT outperformed both prompted and fine-tuned models with significantly larger parameter counts (e.g., 7B). That study supplemented their limited MSH-specific training set of 300 notes with general clinical data from MIMIC-III to improve performance. In contrast, our setting includes a larger, purpose-built corpus of 900 MSH-annotated notes, allowing us to train without relying on external datasets.

##### Implementation Details

We follow a standard classification setup, prepending a “[CLS]” token to each input example. This token is trained to encode task-relevant information and is passed through a feed-forward layer to produce the final binary class probability. Training is conducted with a batch size of 16 and 4 gradient accumulation steps to accommodate longer sequence inputs. We initialize our learning rate as 1e-5.

#### Gemma-2-2b

Gemma-2-2b is a decoder-only, instruction-tuned language model from Google’s Gemma family, designed to deliver strong performance across a wide range of text-based tasks while remaining computationally efficient (Gemma Team, 2024). With 2 billion parameters and a context window of 8192 tokens, it is well-suited for processing long clinical notes without requiring input truncation or windowing. Its relatively compact size makes it easier to fine-tune in constrained environments while still benefiting from the robust pretraining underlying the Gemma model family.

Although Gemma models are typically used in generative applications such as question answering and summarization, we adapt Gemma-2-2b for binary classification. This decision is motivated by prior work, which shows that directly predicting class probabilities from model embeddings outperforms generative approaches that rely on outputting specific tokens (e.g., “true”/”false”), especially in low-data or fine-tuning-constrained scenarios (Hussain et al. 2025).

##### Implementation Details

To repurpose Gemma-2-2b as a classifier, we extract the hidden state of the final token in the input sequence and pass it through a feed-forward layer to produce a scalar probability for the target MSH class. We do not prepend a special classification token such as “[CLS]” because, unlike encoder-based models, decoder-only architectures like Gemma are autoregressive and are not typically pretrained with global-attention tokens. Instead, the final token in the input sequence serves as an effective summary representation, as it attends over all prior tokens in the sequence.

This architecture mirrors the classification setup used for ModernBERT and allows for a direct comparison between encoder and decoder model types. We train Gemma-2-2b with a batch size of 1 and 4 gradient accumulation steps, adjusted for memory constraints associated with its larger parameter count. We additionally use LORA, PEFT, and int-4 quantization. We initialize our learning rate as 2e-5.

### Results

We present our baseline results in Table 3. Overall, the metrics indicate that both models are capable of extracting multidimensional sleep health information from clinical notes. Notably, ModernBERT-large outperforms the findings reported in Hussain et al. (2025), suggesting that our larger, MSH-focused training dataset has improved its classification performance.

Comparing the two models on the NCH-Sleep-Test set, Gemma-2-2b outperforms ModernBERT-large across all metrics. We see Gemma-2-2b has an improvement of +0.18 in F1 and +0.14 improvement in AUROC compared to ModernBERT-large, indicating a superior balance between precision and recall for MSH classification. Additionally, the improvements in both precision and recall suggest that Gemma-2-2b more accurately identifies positive MSH cases while minimizing false positives.

One contributing factor to these performance gains is likely the scale and training paradigm of Gemma-2-2b. As a decoder-only model with 2 billion parameters, it leverages a broader representational capacity to capture complex patterns in unstructured clinical text. In contrast to the 7B parameter models examined in Hussain et al. (2025), which may possess greater initial representational capacity but are more challenging to fine-tune, Gemma-2-2b appears to achieve a better balance of generalizability and fine-tuning flexibility. Meanwhile, ModernBERT-large, though powerful, remains below the peak performance achieved by Gemma-2-2b.

These findings show that, even without explicit expert supervision or keyword-based heuristics, LLMs can learn meaningful representations of MSH from clinical notes. Moreover, they illustrate that model scale and pretraining strategies play critical roles in performance on complex, domain-specific tasks. Consequently, we select Gemma-2-2b as our primary reference point for subsequent experiments that incorporate expert-derived knowledge, with ModernBERT-large serving as a strong yet more resource-efficient alternative.

## Experiment 2: Oracle Classification

As part of the annotation process for the NCH-Sleep dataset, expert annotators identified specific spans within each clinical note that supported their labeling decisions for positive instances of MSH classes. We refer to these as oracle spans: segments of text that highlight the rationale behind a human expert’s classification, but which would not be available to a model during real-world inference. These spans represent what experts deemed the most relevant evidence for each class and thus serve as a proxy for gold-standard interpretive signals.

In this experiment, we investigate whether training a model solely on these oracle spans improves or impairs classification performance. On one hand, focusing the model’s attention on highly relevant expert-identified content might enhance learning by eliminating distracting or irrelevant context. On the other hand, these spans may rely on implicit clinical reasoning or background knowledge that is not self-contained within the text. In such cases, restricting input to oracle spans may obscure necessary context and reduce model effectiveness.

Our central question is whether oracle-guided training can help the model recover or exceed baseline performance, or whether it reveals a mismatch between how human experts reason about clinical evidence and how models derive signal from raw text. Improved performance would suggest that LLMs benefit from explicit supervision in the form of targeted evidence. Conversely, reduced performance may indicate that expert annotations omit crucial surrounding context, or that models rely on broader cues not captured by span-level annotations.

### Methods

For each note, we have expert-annotated spans linked to one or more of the nine MSH classes. Because our classification task treats each MSH class independently, incorporating only the oracle spans associated with the target class would allow the model to trivially associate the presence of any oracle span with a positive classification. To avoid this, we aggregate the oracle spans from all MSH classes within each note. Then, for each binary MSH classification example, we provide the complete set of oracle spans. In this manner, the model is still directed toward the expert-identified information without being explicitly told which class should be positively classified.

We consider two primary approaches for incorporating oracle spans: focusing model attention within the note, Oracle_Anno, or removing irrelevant context from the note, Oracle_Only. For both cases, if spans overlap, they are combined into one larger contiguous span.

#### oracle_anno

In this case, we seek to avoid removing any information from the note but rather provide signals which may guide the model to focus on particular sections of the note.

To construct the context string, we identify each oracle_span present in the note and prepend/append it with “<<”/”>>”. These tokens were selected for their simplicity and because neither “<” or “>” occurs within NCH-Sleep, minimizing the chance of interference with content within the note. Outside of the added tokens, the rest of the note remains identical.

#### oracle_only

In this case, we remove content from the note, leaving behind only the oracle spans. Contrasted to oracle_anno, this enforces rather than guides the model’s attention so that decisions cannot be made based on any other part of the note. We construct the context string by concatenating each oracle_span present within the note with a ‘[SEP]’ token joining them, ordered according to the position in the note.

~~~
context_string = “oracle_span_1 [SEP] oracle_span_2 [SEP] … oracle_span_n”
~~~

#### Implementation Details

We fine-tune the top-performing baseline configuration from Experiment 1, Gemma-2-2b (retaining all noted hyperparameters from Experiment 1). All other training environment and implementation details are retained from Experiment 1. When we apply oracle_anno or oracle_only, we do so for both the training and test set.

### Results

In Table 4, we present the results from our oracle-guided experiments alongside the baseline. Overall, incorporating expert-annotated spans into the model input markedly improves performance on the MSH classification task. The Oracle_Anno approach, where the full clinical note is preserved with expert-identified spans marked, yields notable gains: +0.10 in F1 score, and +0.08 in AUROC. These improvements suggest that explicitly highlighting key segments within the broader context helps the model to more effectively focus on clinically relevant information.

**Table 4:** Results for our oracle models trained on NCH-Sleep-Train and evaluated on NCH-Sleep-Test. Values presented are means across 5 training trials with different seeds. Standard deviations are presented within parentheses.

| Model | Precision | Recall | F1 | AUROC |
| --- | --- | --- | --- | --- |
| Baseline | 0.53 (0.13) | 0.77 (0.11) | 0.62 (0.13) | 0.86 (0.08) |
| Oracle_Anno | 0.63 (0.04) | 0.86 (0.05) | 0.73 (0.04) | 0.93 (0.02) |
| Oracle_Only | 0.74 (0.05) | 0.92 (0.02) | 0.82 (0.03) | 0.97 (0.00) |

The Oracle_Only configuration, which restricts input exclusively to the expert-annotated spans, produces even more pronounced gains: +0.20 in F1 score, and +0.11 in AUROC. This sharp improvement highlights the discriminative value of the expert-identified spans alone, suggesting that models can achieve even higher performance when irrelevant or noisy content is removed entirely. By stripping away unrelated text, Oracle_Only likely reduces distraction and helps the model focus solely on high-signal content, effectively increasing the signal-to-noise ratio during both training and inference.

Together, these findings show that integrating expert guidance via oracle spans substantially enhances the classification of MSH from clinical notes. The enhancements in F1 and AUROC demonstrate that combining domain expertise with data-driven methods can bridge the gap between raw text representations and the nuanced clinical insights necessary for accurate MSH classification.

## Experiment 3: Keyword-Based Classification

While Experiment 2 demonstrated the substantial value of oracle spans for improving MSH classification, this form of supervision represents an idealized scenario. In practical settings, expert-generated span annotations are not available at inference time. Thus, we explore whether a more feasible form of expert signal—domain-informed keywords—can serve as an effective proxy. Specifically, we examine whether keyword-guided context can enhance classification by nudging models toward clinically relevant segments of the note without requiring detailed annotation.

The NCH-Sleep dataset was initially constructed by gathering clinical notes that contain at least one of a curated list of 348 sleep-related keywords developed by clinical sleep experts. This keyword list expands upon prior research and encompasses terms relevant to behaviors, symptoms, medications, and other sleep health dimensions identified by surveying sleep-related ontologies, discovering synonymous terms and abbreviations found in NCH notes, and interviews with clinicians. These terms serve as domain-informed indicators that signal the potential presence of MSH-relevant content. Our goal in this experiment is to determine whether leveraging these keywords to structure model input can replicate some of the benefits of oracle supervision while remaining applicable to real-world deployment.

### Methods

As in Experiment 2, we evaluate two approaches for incorporating keyword information into the input context: keyword_anno, which highlights keywords within the full note, and keyword_only, which restricts model input to localized windows surrounding keyword occurrences.

Because each note in NCH-Sleep was originally retrieved based on keyword presence, every note contains at least one keyword. We identify keyword matches using case-insensitive string matching, but enforce whole-word boundaries to avoid unintended subword matches (e.g., the keyword “hyper” would not match “hyperactivity,” but would match “hyper!”).

#### keyword_anno

For each keyword found in a note, we mark it by wrapping the word with “<<” and “>>”. These special tokens are used to signal the keyword’s position within the note without altering the underlying content or removing context. This method mirrors the oracle_anno approach from Experiment 2, guiding the model’s attention while preserving the full narrative structure of the note.

#### keyword_only

In this setting, we extract fixed-length windows of text around each keyword. For each match, we include X characters before and after the keyword, forming a localized keyword span. These spans are concatenated in order of appearance, separated by “[SEP]” tokens, to form the new context string. All other parts of the note are removed. This approach is analogous to oracle_only, but the guiding signal is a heuristic rather than a manually annotated span.

We experiment with nine window sizes for X ∈ {0, 10, 20, 40, 80, 160, 320, 640, 1280}, allowing us to evaluate how much surrounding context is necessary to preserve performance. If a window boundary would cut through a word, we extend it to the nearest whitespace to preserve word integrity.

#### Implementation Details

We continue to fine-tune the Gemma-2-2b model with all hyperparameters and training settings retained from Experiments 1 and 2. As with oracle-based approaches, the keyword transformations are applied consistently across both training and test sets.

### Results

We present in Table 5 the results of our keyword-guided classification experiments. Overall, the findings confirm that expert-curated keywords can meaningfully improve model performance on the MSH classification task, even in the absence of detailed span-level supervision.

**Table 5:** Results for our keyword models trained on NCH-Sleep-Train and evaluated on NCH-Sleep-Test. Values presented are means across 5 training trials with different seeds. Standard deviations are presented within parentheses.

| Task | Precision | Recall | F1 | AUROC |
| --- | --- | --- | --- | --- |
| Baseline | 0.53 (0.13) | 0.77 (0.11) | 0.62 (0.13) | 0.86 (0.08) |
| keyword_anno | 0.58 (0.06) | 0.79 (0.03) | 0.67 (0.05) | 0.90 (0.02) |
| keyword_only_0 | 0.59 (0.06) | 0.74 (0.04) | 0.66 (0.02) | 0.89 (0.01) |
| keyword_only_10 | 0.58 (0.05) | 0.76 (0.03) | 0.65 (0.04) | 0.89 (0.02) |
| keyword_only_20 | 0.62 (0.05) | 0.81 (0.08) | 0.70 (0.06) | 0.91 (0.03) |
| keyword_only_40 | 0.65 (0.09) | 0.81 (0.05) | 0.72 (0.07) | 0.92 (0.03) |
| keyword_only_80 | 0.60 (0.07) | 0.82 (0.02) | 0.69 (0.04) | 0.91 (0.01) |
| keyword_only_160 | 0.55 (0.08) | 0.83 (0.03) | 0.66 (0.06) | 0.90 (0.03) |
| keyword_only_320 | 0.51 (0.09) | 0.80 (0.05) | 0.62 (0.08) | 0.87 (0.05) |
| keyword_only_640 | 0.66 (0.07) | 0.87 (0.01) | 0.75 (0.05) | 0.94 (0.02) |
| keyword_only_1280 | 0.54 (0.08) | 0.73 (0.13) | 0.62 (0.10) | 0.86 (0.07) |

The keyword_anno condition, where keywords are highlighted within the full clinical note, yields a modest improvement over the baseline: a +0.05 gain in F1 score and +0.04 gain in AUROC. These gains suggest that marking keyword locations helps the model attend to clinically relevant regions without removing the broader context. However, since the model still has access to the entire note, its predictions may be influenced by irrelevant content, limiting interpretability. This contrasts with conditions where the context is explicitly constrained, offering clearer insight into what information drove the model’s decision.

In the keyword_only condition, where input is restricted to fixed-length windows around each keyword, performance varies with window size. Very short windows (e.g., 0–20 characters) limit recall but improve precision, while very large windows (e.g., 1280 characters) dilute the keyword signal and begin to approximate the full-note setting. The best-performing configuration is keyword_only_640, which achieves a +0.13 increase in F1 and a +0.08 gain in AUROC over the baseline. These results highlight the importance of appropriately tuning context length to balance informativeness and noise.

Importantly, even smaller windows, such as keyword_only_40, which match or slightly outperform the baseline, present a compelling opportunity for clinical application. By retaining high predictive performance within a much smaller input scope, these configurations support more interpretable outputs. Given that the average clinical note in NCH-Sleep is nearly 6000 characters long, reducing the model’s input to a few hundred targeted characters can help surface concise, high-yield evidence. This creates the potential for human-in-the-loop decision-making, where clinicians or researchers reviewing an alert can quickly scan the exact information that the model used to arrive at a prediction. Such a design could mitigate alert fatigue and foster trust in automated systems, allowing expert intuition to complement model outputs in real-time workflows.

However, it is important to note that the performance of keyword-guided models may be inflated due to selection bias in the dataset. Since all notes in NCH-Sleep were originally retrieved based on the presence of expert-derived keywords, the test set is inherently enriched for examples that align with the keyword set. As a result, clinical descriptions of MSH that fall outside the predefined vocabulary may be underrepresented or entirely absent. This limitation suggests that keyword-only models may not generalize as well to unseen or more diverse documentation styles, where relevant mentions are phrased differently or implicitly described.

In sum, keyword-based supervision offers a scalable and practical middle ground between baseline free-text classification and oracle-guided annotation. While it does not reach the performance ceiling of oracle-based approaches, it meaningfully narrows the gap and introduces opportunities for improved interpretability and clinician engagement in decision support contexts.

## Experiment 4: Keyword Overlap with Oracle

Experiments 2 and 3 demonstrated that expert-derived signals, both oracle spans and curated keyword lists, can significantly improve model performance on MSH classification. While keyword-based methods offer a scalable and realistic alternative to manually annotated spans, they do not fully close the performance gap. This raises an important question: to what extent do curated keywords capture the same information as oracle spans? Understanding this alignment is essential for evaluating the precision and recall characteristics of keyword-based supervision and for informing the design of future heuristic guidance strategies.

In this experiment, we quantify the overlap between keywords and oracle spans in the NCH-Sleep dataset to assess whether the observed performance differences stem from limitations in keyword coverage, specificity, or both. Specifically, we aim to understand whether keywords tend to co-occur with oracle spans, whether they occur frequently outside them, and how individual keywords vary in informativeness.

### Methods

We perform the following analyses to characterize the relationship between keywords and oracle spans. All evaluations were done on the training split of NCH-Sleep:

#### Oracle w/ Keyword

What proportion of oracle spans contain at least one keyword match? This gives us a sense of how often the keyword list successfully targets expert-annotated evidence.

#### Keyword w/o Oracle

What proportion of keyword matches in a note occur within an oracle span? This reflects the precision of keywords (i.e., how often they align with expert-identified text rather than appearing elsewhere in the note).

#### Mutual Information Analysis

To go beyond aggregate overlap statistics, we also compute a per-keyword informativeness score. For each keyword, we calculate the proportion of its appearances that fall inside oracle spans versus outside them. This yields a mutual information-like signal: a score of 1 indicates the keyword always appears in oracle evidence, whereas a score of 0 means it never does (Equation 1). This analysis helps identify which keywords are most reliably tied to expert-classified content and which may contribute to noise in keyword-only input representations.

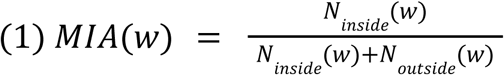

### Results

Across the dataset, we find that 70.90% of oracle spans contain at least one keyword, suggesting that the curated keyword list successfully captures a majority of expert-identified evidence. However, this also implies that nearly 30% of oracle-annotated spans contain no keyword at all, highlighting a notable recall gap in the keyword list. These “missed” spans likely reflect more implicit or paraphrased clinical descriptions that are not easily captured by lexical heuristics alone.

On the precision side, 69.92% of all keyword matches occur within oracle spans, meaning roughly 30% of keyword appearances fall outside the expert-highlighted regions. This indicates that while keywords tend to align with relevant content, a nontrivial portion may appear in more generic or less diagnostic portions of the note, potentially introducing noise when used to guide model input.

To further characterize keyword informativeness, we compute a Mutual Information Approximation (MIA) score for each keyword, defined as the proportion of its appearances that fall within oracle spans (Table 6). A score of 1 indicates perfect alignment with oracle spans, whereas a score of 0 means the keyword never appears in expert-labeled evidence. Among the 348 total keywords, only 156 (44.83%) appear in the dataset at all, indicating that over half of the keyword list is either overly rare or mismatched with the linguistic patterns in the NCH-Sleep notes.

**Table 6:** Top 5 (by MIA), Most Common 5 (by Note Freq), and Bottom 5 (by MIA) keywords.

| Keyword | note_freq(%) | MIA |
| --- | --- | --- |
| difficulty sleeping | 5.25 | 1 |
| trouble falling asleep | 3.5 | 1 |
| difficulty falling asleep | 3.08 | 1 |
| naps | 3 | 1 |
| poor sleep | 2.67 | 1 |
| sleep | 73.92 | 0.92 |
| bedtime | 27.75 | 0.336 |
| wheezing | 26 | 0 |
| melatonin | 21.42 | 0.992 |
| sleeping | 20.17 | 0.938 |
| stops breathing at night | 0.08 | 0 |
| eats late | 0.08 | 0 |
| unable to sleep | 0.08 | 0 |
| grinds teeth | 0.08 | 0 |
| lavender | 0.08 | 0 |

The mean MIA for keywords that appear in the dataset is 0.738, with a standard deviation of 0.380. This high variance suggests a wide range in keyword quality. Some keywords consistently align with oracle spans and serve as strong signals (e.g., “melatonin”, “poor sleep”, “trouble falling asleep”), while others occur frequently outside oracle evidence or not at all (e.g., “wheezing”, “eats late”, “lavender”), thereby introducing noise when used indiscriminately. Notably, several of the most frequent keywords, such as “bedtime” and “sleep”, exhibit middling or low MIA scores, underscoring that frequency does not necessarily correlate with precision.

These findings reveal a mixed but informative picture of keyword coverage. First, the keyword list demonstrates high recall: keywords appear in over 70% of oracle spans, indicating that the list successfully captures many relevant concepts flagged by experts. However, the precision is more moderate; approximately 30% of keyword matches occur outside oracle spans, suggesting that not all keyword occurrences are informative and that some may introduce noise into the model’s input. Finally, there is wide variability in keyword quality. Some keywords are highly discriminative and consistently align with oracle spans, while others are either too generic, overly frequent, or entirely irrelevant to the annotated evidence.

This analysis provides a clearer understanding of why keyword-guided models perform well, but not as well as oracle-guided ones. While keyword-based approaches offer a scalable and interpretable alternative to manual annotation, their effectiveness is bounded by the coverage and specificity of the underlying keyword list. Closing this gap will require developing more context-sensitive heuristics, such as dynamic keyword expansion, concept embeddings, or phrase-level matching, to capture latent or implied clinical information that static keywords miss.

## Experiment 5: Bias Analysis

Having a comprehensive understanding of model bias is critical for any system that is oriented towards clinical application. To evaluate the effects of bias in our best performing systems, we stratify performance by demographics, department, and class. This evaluation will indicate the limitations of these systems as they interact with various facets of real-world application.

### Methods

We evaluate a subset of configurations from experiments 1, 2, and 3. Namely, we consider the following configurations: Baseline, Oracle_only, keyword_only_40, and keyword_anno.

Baseline, keyword_only_40, and keyword_anno represent possible real-life implementations, while oracle_only presents an expert-mediated idealized scenario.

#### Demographic Stratification

To investigate potential disparities in data distribution and model accuracy across population subgroups, we analyze the performance of our top-performing models separately by gender and by race.

#### Insurance Stratification

Type of insurance may serve as a proxy for healthcare access and, in turn, influence how MSH-related information is presented and recorded. We examine model behavior across groups with private insurance, public insurance, or both types.

#### Department Stratification

Since documentation practices and EHR formatting can differ significantly across clinical departments, we assess model performance stratified by both department and gender to understand how such variations may affect outcomes.

#### Performance by Class

Some MSH categories may pose greater challenges for classification than others, particularly depending on the distribution of training examples. We report performance metrics across each class to identify these differences.

### Results

We present our bias analysis results across demographic, insurance, departmental, and class-based stratifications in Tables 7 through 11. Overall, our analyses indicate strong, stable model performance across the evaluated subgroups, with a few notable exceptions that highlight areas for targeted improvements.

When examining performance stratified by patient gender (Table 7), we observed nearly identical performance between female (n=140) and male (n=160) subgroups across all evaluated configurations. The AUROC scores consistently ranged between 0.86–0.97, with minimal differences (<0.01) between genders, suggesting that our models do not display significant gender bias in MSH classification.

**Table 7:** Mean (5 trials) AUROC over NCH-Sleep Test for select models stratified by patient gender. Standard deviations are parenthesized. n = XX refers to note count.

| model | female (n=140) | male (n=160) |
| --- | --- | --- |
| baseline | 0.86 (0.08) | 0.86 (0.09) |
| keyword_anno | 0.90 (0.03) | 0.90 (0.02) |
| keyword_only_40 | 0.92 (0.03) | 0.92 (0.03) |
| oracle_only | 0.96 (0.00) | 0.97 (0.00) |

In contrast, stratification by patient race (Table 8) revealed modest disparities. While the baseline and keyword-guided models maintained comparable performance (AUROC ∼0.84–0.92) across African American (n=96), Latino (n=40), Multi-racial (n=24), and White (n=128) groups, performance dropped notably for the “Other” category (n=12), composed primarily of smaller representation groups (African, Asian, Indigenous). Here, keyword-guided models especially suffered, dipping as low as 0.73 AUROC in the keyword_anno setting.

**Table 8:**
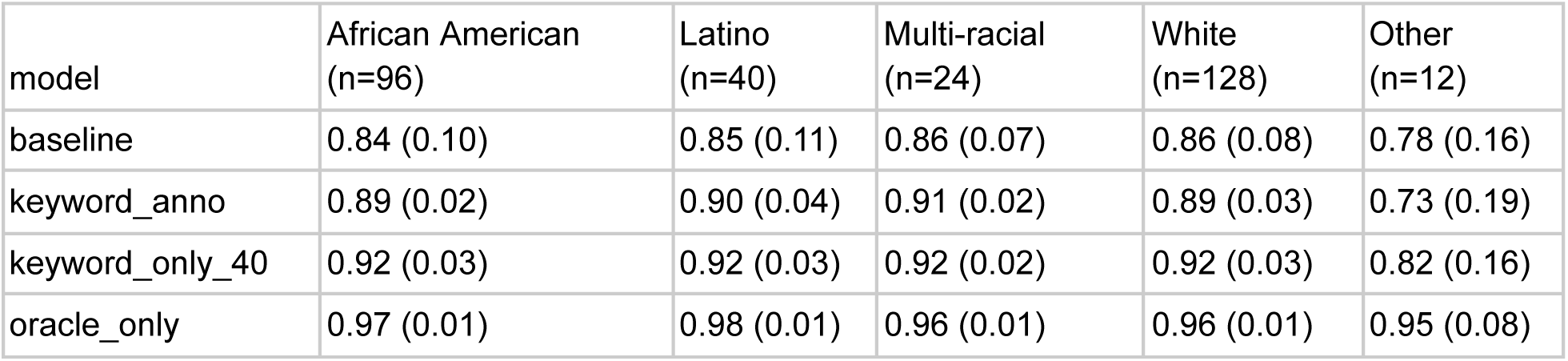
Mean (5 trials) AUROC over NCH-Sleep Test for select models stratified by patient race. Standard deviations are parenthesized. n = XX refers to note count. Other category contains African (n=9), Asian (n=2), and Indigenous (n=1)

| model | African American (n=96) | Latino (n=40) | Multi-racial (n=24) | White (n=128) | Other (n=12) |
| --- | --- | --- | --- | --- | --- |
| baseline | 0.84 (0.10) | 0.85 (0.11) | 0.86 (0.07) | 0.86 (0.08) | 0.78 (0.16) |
| keyword_anno | 0.89 (0.02) | 0.90 (0.04) | 0.91 (0.02) | 0.89 (0.03) | 0.73 (0.19) |
| keyword_only_40 | 0.92 (0.03) | 0.92 (0.03) | 0.92 (0.02) | 0.92 (0.03) | 0.82 (0.16) |
| oracle_only | 0.97 (0.01) | 0.98 (0.01) | 0.96 (0.01) | 0.96 (0.01) | 0.95 (0.08) |

Interestingly, the oracle_only configuration largely mitigated this disparity, with the AUROC recovering to 0.95, suggesting that explicit expert guidance can significantly counteract biases introduced by small sample sizes or underrepresentation. This finding highlights the importance of expert mediation for model fairness in underrepresented groups.

The type of insurance was evaluated as a proxy for healthcare access disparities (Table 9). All models generally exhibited stable AUROC scores (0.85–0.98) across patients with public insurance (n=199), private insurance (n=48), and combined public/private coverage (n=51). The keyword_only_40 configuration notably performed highest across the combined insurance group (AUROC=0.93), while maintaining a strong performance for purely public or private groups. Although slight performance fluctuations were observed, no significant bias pattern emerged based on insurance type, indicating robustness of our models across varying levels of healthcare accessibility.

**Table 9:** Mean (5 trials) AUROC over NCH-Sleep Test for select models stratified by patient insurance. Standard deviations are parenthesized. n = XX refers to note count.

| model | Public (n=199) | Public/Private (n=51) | Private (n=48) | Other (n=2) |
| --- | --- | --- | --- | --- |
| baseline | 0.85 (0.09) | 0.87 (0.08) | 0.86 (0.06) | 0.82 (0.14) |
| keyword_anno | 0.90 (0.03) | 0.91 (0.03) | 0.87 (0.02) | 0.86 (0.08) |
| keyword_only_40 | 0.92 (0.03) | 0.93 (0.03) | 0.90 (0.03) | 0.90 (0.07) |
| oracle_only | 0.96 (0.00) | 0.98 (0.00) | 0.96 (0.01) | 0.97 (0.02) |

Given differences in documentation standards and clinical norms, performance was stratified by originating clinical departments (Table 10). Baseline performance was fairly consistent (AUROC ∼0.85–0.87) across Primary Care (n=174), Behavioral Health (n=106), and Other departments (Healthy Weight and School Based, n=20). Keyword-guided models slightly improved performance (AUROC ∼0.88–0.96), with keyword_only_40 performing exceptionally well for notes originating in the smaller department categories (AUROC=0.96). Oracle_only remained the strongest across all departments (AUROC=0.96–0.99), suggesting that expert annotations robustly generalize across clinical documentation practices, providing valuable insight for future deployment in diverse departmental contexts.

**Table 10:** Mean (5 trials) AUROC over NCH-Sleep Test for select models stratified by note department. Standard deviations are parenthesized. n = XX refers to note count. Other category contains Healthy Weight (n=14) and School Based (n=6)

| model | Primary Care (n=174) | Behavioral Health (n=106) | Other (n=20) |
| --- | --- | --- | --- |
| baseline | 0.86 (0.09) | 0.85 (0.08) | 0.87 (0.11) |
| keyword_anno | 0.91 (0.02) | 0.88 (0.03) | 0.94 (0.04) |
| keyword_only_40 | 0.92 (0.03) | 0.91 (0.04) | 0.96 (0.03) |
| oracle_only | 0.96 (0.00) | 0.97 (0.00) | 0.99 (0.00) |

We further investigated class-level performance variations (Table 11). Baseline performance exhibited variability, with AUROC scores ranging from 0.81 (”Behavior”) to 0.91 (”Timing”). Keyword-based approaches significantly boosted performance for classes such as “Alertness,” “Duration,” “Medication,” and “Timing,” achieving AUROCs in excess of 0.95 in keyword_only_40. However, classes like “Behavior,” “Efficiency,” and “Intervention” demonstrated lower sensitivity to keyword-only guidance, suggesting either insufficient keyword coverage or increased complexity in their linguistic representation. Oracle_only substantially improved classification across all classes, suggesting that explicit expert annotations remain superior for consistently capturing nuanced clinical phenomena.

**Table 11:** Mean (5 trials) AUROC over NCH-Sleep Test for select models stratified by MSH Class. Standard deviations are parenthesized.

| Task | Alertness | Behavior | Duration | Efficiency | Satisfaction | Disorder | Intervention | Medication | Timing |
| --- | --- | --- | --- | --- | --- | --- | --- | --- | --- |
| baseline | 0.85<br>(0.08) | 0.81<br>(0.08) | 0.87<br>(0.11) | 0.82<br>(0.06) | 0.84<br>(0.08) | 0.83<br>(0.12) | 0.84<br>(0.08) | 0.90<br>(0.09) | 0.91<br>(0.09) |
| keyword_anno | 0.90<br>(0.04) | 0.82<br>(0.02) | 0.92<br>(0.06) | 0.86<br>(0.03) | 0.84<br>(0.02) | 0.92<br>(0.04) | 0.85<br>(0.01) | 0.98<br>(0.01) | 0.95<br>(0.04) |
| keyword_only_40 | 0.96<br>(0.01) | 0.87<br>(0.02) | 0.94<br>(0.02) | 0.86<br>(0.01) | 0.91<br>(0.05) | 0.92<br>(0.06) | 0.88<br>(0.03) | 0.95<br>(0.02) | 0.96<br>(0.02) |
| oracle_only | 0.98<br>(0.01) | 0.94<br>(0.02) | 0.98<br>(0.00) | 0.88<br>(0.01) | 0.98<br>(0.00) | 0.98<br>(0.00) | 0.94<br>(0.01) | 0.99<br>(0.00) | 0.99<br>(0.00) |

Our bias analysis demonstrates that while our models perform robustly across most subgroups and classes, certain areas warrant further attention. Specifically, targeted efforts are needed to improve model performance for underrepresented racial groups, and to enhance keyword coverage or develop more nuanced heuristics for challenging MSH classes such as “Behavior,” “Efficiency,” and “Intervention.” These results underscore the continued importance of incorporating expert oversight, particularly for addressing model biases and ensuring equitable application across diverse clinical populations and contexts.

## Discussion

In this work, we explored multiple strategies for classifying multidimensional sleep health (MSH) within electronic health record (EHR) text using large language models (LLMs). Our baseline experiment (Experiment 1) established that even with minimal supervision (e.g. the raw clinical note and a brief description of the target class) models such as Gemma-2-2b and ModernBERT-large can learn significant aspects of MSH. However, both precision and recall remained imperfect, underlining the complexity of capturing nuanced sleep-related phenomena solely from raw text.

To address this limitation, Experiment 2 and Experiment 3 introduced two forms of expert-driven guidance: oracle spans (expert-annotated textual evidence) and curated keywords, respectively. In Experiment 2, restricting or highlighting the model’s attention to oracle spans yielded the highest gains in performance. Specifically, the Oracle_Only configuration demonstrated that by removing irrelevant or noisy text, models could pinpoint clinically meaningful indicators of sleep health far more effectively. This result underscores how human expertise can substantially refine model attention, effectively eliminating ambiguity arising from extraneous EHR content. However, such an approach requires continuous human annotation, which is resource-intensive and not universally feasible.

Experiment 3 sought a more practical balance between fully manual annotation and baseline free-text classification through a large, expert-curated keyword list. Our results showed that keyword-based transformations indeed improved classification metrics, although they did not match the performance ceiling achieved with oracle spans. Furthermore, experiments with different window sizes around these keywords revealed a trade-off between capturing sufficient context and maintaining a focused, interpretable input. Notably, while large windows or full-note contexts may maximize performance, smaller windows (e.g., keyword_only_40) retained strong classification metrics and offered improved interpretability. This smaller, more digestible context can facilitate human-in-the-loop reviews, enabling clinicians or researchers to quickly examine the precise segments of the note used by the model. Such transparency may improve trust in automated systems and allow for real-time clinical judgment, bias detection, or other contextual assessments that are crucial in patient care environments.

Experiment 4’s keyword-oracle overlap analysis demonstrated that while keywords captured most expert-labeled evidence, they also missed a nontrivial fraction of spans. Inversely, a substantial portion of keyword occurrences fell outside oracle spans, thereby potentially introducing noise. These findings clarify why keyword-only strategies can enhance performance but still lag behind oracle-based approaches: the coverage and specificity of the keyword list are incomplete. This limitation motivates future work in dynamic, context-sensitive keyword expansion or the integration of broader NLP heuristics (e.g., phrase embeddings, topic modeling) to better approximate the granularity and context-awareness of expert annotations.

Finally, Experiment 5 assessed how these models perform across different patient demographics, departments, insurance types, and MSH classes. The results revealed fairly consistent outcomes across most subgroups but highlighted potential biases, particularly for underrepresented racial categories and certain MSH classes (e.g., “Behavior,” “Efficiency,” and “Intervention”). The improved fairness under oracle_only conditions suggests that more targeted or inclusive expert involvement can help mitigate biases stemming from uneven data representation.

Collectively, these experiments highlight that LLMs are capable of implicitly learning where to focus within clinical notes, achieving strong baseline performance without explicit supervision. However, the substantial performance gains observed with oracle annotations suggest that expert-guided signals still offer untapped value. Keyword-based methods helped close part of this gap by anchoring model attention to known high-signal terms, and they introduced the additional benefit of interpretability, particularly when smaller context windows were used. Still, these methods come with trade-offs: coverage is incomplete, some matches are noisy, and they lack the precision of expert annotations. Future work should explore ways to enhance heuristic span identification, such as training named entity recognition (NER) models to expand keyword sets or directly predict oracle-like spans. These strategies could further bridge the gap between expert-level supervision and scalable, interpretable clinical NLP systems.

## Conclusion

Our comprehensive evaluation reveals that domain-informed strategies, ranging from oracle spans to heuristic keywords, can significantly boost MSH classification performance beyond what is achievable with baseline, free-text inputs. While manually highlighted spans produce the most substantial improvements, expert-curated keyword approaches represent a more feasible solution for day-to-day clinical practice, narrowing the gap at a lower cost in human effort.

Moreover, smaller-window or keyword-focused approaches (e.g., keyword_only_40) may be especially valuable in settings that require a human-in-the-loop workflow. By constraining the model’s attention to concise, high-yield text, these methods preserve competitive predictive performance and increase interpretability, thereby promoting clinician trust and enabling swift expert review. In real-world scenarios, balancing performance with transparency and fairness is paramount; our results suggest that iterative refinements to both keyword coverage and domain-focused heuristics, combined with selective expert oversight, can deliver robust, interpretable pipelines for MSH detection at scale.

## Data Availability

The data for this work will not be available

## Appendix

## A.1 Binary vs Multi-class Classification

In our primary experiments, we frame MSH classification as a set of conditionally independent binary classification tasks: one for each of the nine MSH categories. This approach treats each class independently, allowing us to formulate a simple and flexible task structure where a model is queried for the presence of each class individually. Practically, this means the model receives a clinical note along with a class description and predicts a binary outcome (present or not present) for that specific class. The independence assumption simplifies modeling and supports modularity, but it prevents the model from learning correlations between classes (e.g., the co-occurrence of sleep disorders and sleep medication).

In this section, we evaluate the impact of relaxing this assumption by reframing the task as multi-label classification, where the model simultaneously predicts the presence or absence of all nine MSH classes. This formulation allows for inter-class dependencies to be captured through shared model parameters and hidden representations. Our goal is to assess whether this joint prediction approach improves classification performance and better reflects the structure of real-world clinical documentation, where mentions of different sleep health indicators often co-occur or influence each other.

### Method

#### Models

We compare ModernBERT-large and Gemma-2-2b. Rather than our feedforward layer for classification having 1 output (positive class presence probability), it now has 9 outputs (1 for each MSH class). Since we assume that multiple classes can be true at once, we continue to use Binary Cross Entropy (BCE) loss.

#### Task Definition

Since we do not consider only one class at a time, we remove the class_name and class_description from our input. Likewise, we expand our binary prediction to instead create a vector of 9 binary predictions, one for each MSH class. For this same reason, where our original test set represented 2700 comparisons (300 notes x 9 classes), we now conduct a smaller 300 comparisons,

~~~
Original_Input: {‘clinical_note’: {clinical_note}, ‘class_label’: {class_name}, ‘class_description’: {class_description}}
Original_Output: 1/0
MultiClass_Input: {‘clinical_note’: {clinical_note}}
MultiClass_Output: [0/1, 0/1, 0/1, 0/1, 0/1, 0/1, 0/1, 0/1, 0/1]
~~~

### Results

Table A.1 presents the per-class and overall AUROC scores for both the binary and multi-label (multi-class) configurations using ModernBERT-large and Gemma-2-2b. Overall, we observe that the binary classification setup consistently outperforms the multi-label alternative across nearly all classes, with the largest margins appearing in the Gemma-2-2b configuration.

Gemma-2-2b in the binary setting achieves the highest overall AUROC (0.86), outperforming its multi-label counterpart by 0.14 points. Notably, classes like ‘Alertness’, ‘Duration’, and ‘Timing’ benefit the most from the binary framing, each showing improvements of at least 0.15 AUROC. This suggests that treating classes independently allows the model to better specialize for individual classification subtasks, possibly due to the focused nature of each training example and the additional context provided via class descriptions.

ModernBERT-large exhibits similar trends, although the difference between binary and multi-label performance is more muted. Both configurations hover around an overall AUROC of 0.71–0.72, with modest variation across classes. This suggests that for smaller models or lower-capacity settings, the benefit of class-specific contextualization (via binary prompts) may be less pronounced.

The reduced performance of multi-label configurations likely stems from two key limitations:

1. Loss of class-specific context: Removing the class label and description deprives the model of helpful guidance about the task it is solving, particularly for subtle or sparsely annotated MSH categories.
2. Joint prediction interference: Simultaneously predicting all nine labels may introduce optimization challenges, especially when class imbalance is high and class correlations are non-trivial but poorly captured.

While multi-label classification offers the advantage of efficiency (i.e., one pass per note vs. nine), our findings suggest that this comes at a cost to performance. Especially in clinically sensitive applications, where recall and precision are critical, binary classification offers more reliable outcomes.

In sum, while multi-label models may be attractive for inference speed or deployment simplicity, binary classification remains the preferred choice for accuracy and robustness in MSH classification, particularly when high-performing and interpretable systems are the goal. Future work may explore hybrid models that combine the benefits of multi-label efficiency with explicit task conditioning (e.g., by reintroducing class descriptors or learned label embeddings).

**Table A.1:**
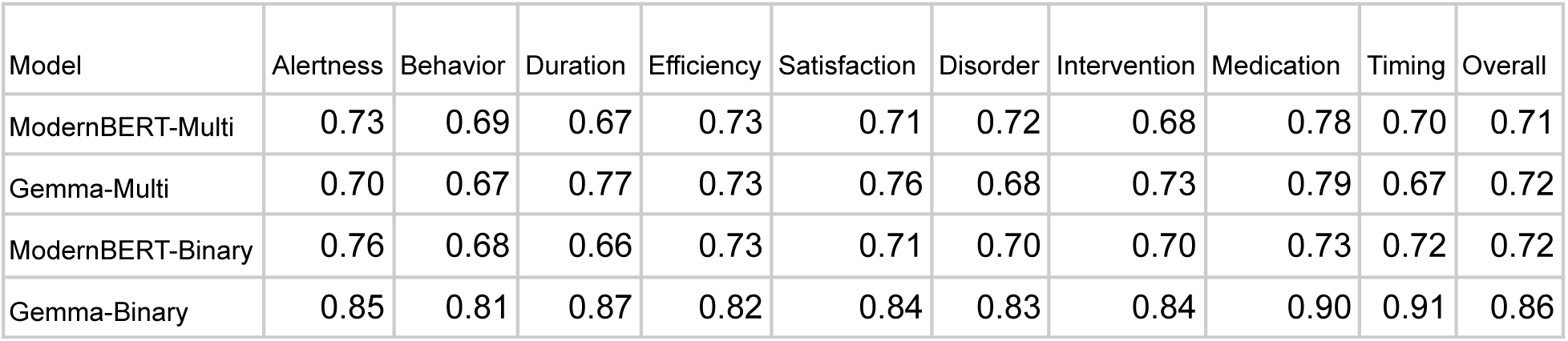
AUROC scores, per class and overall, for binary and multi-class classifiers using both ModernBERT-large and Gemma-2-2b.

| Model | Alertness | Behavior | Duration | Efficiency | Satisfaction | Disorder | Intervention | Medication | Timing | Overall |
| --- | --- | --- | --- | --- | --- | --- | --- | --- | --- | --- |
| ModernBERT-Multi | 0.73 | 0.69 | 0.67 | 0.73 | 0.71 | 0.72 | 0.68 | 0.78 | 0.70 | 0.71 |
| Gemma-Multi | 0.70 | 0.67 | 0.77 | 0.73 | 0.76 | 0.68 | 0.73 | 0.79 | 0.67 | 0.72 |
| ModernBERT-Binary | 0.76 | 0.68 | 0.66 | 0.73 | 0.71 | 0.70 | 0.70 | 0.73 | 0.72 | 0.72 |
| Gemma-Binary | 0.85 | 0.81 | 0.87 | 0.82 | 0.84 | 0.83 | 0.84 | 0.90 | 0.91 | 0.86 |

## References

[1] Benjamin Warner, Antoine Chaffin, Benjamin Clavié, Orion Weller, Oskar Hallström, Said Taghadouini, Alexis Gallagher, Raja Biswas, Faisal Ladhak, Tom Aarsen, Nathan Cooper, Griffin Adams, Jeremy Howard, & Iacopo Poli. (2024). Smarter, Better, Faster, Longer: A Modern Bidirectional Encoder for Fast, Memory Efficient, and Long Context Finetuning and Inference.

[2] Edward J. Hu, Yelong Shen, Phillip Wallis, Zeyuan Allen-Zhu, Yuanzhi Li, Shean Wang, Lu Wang, & Weizhu Chen. (2021). LoRA: Low-Rank Adaptation of Large Language Models.

[3] Sourab Mangrulkar, Sylvain Gugger, Lysandre Debut, Younes Belkada, Sayak Paul, & Benjamin Bossan. (2022). PEFT: State-of-the-art Parameter-Efficient Fine-Tuning methods.

[4] Horner, Matthew, Noor Abul-el-rub, Mary Mays, and Diego Mazzotti. 2022. “0610 Development of a Rule-Based Text Mining Algorithm to Identify Sleep Complaints in Primary Care Progress Notes.” Sleep 45 (Supplement_1): A267–68. 10.1093/sleep/zsac079.607.

[5] Sivarajkumar, Sonish, Thomas Yu Chow Tam, Haneef Ahamed Mohammad, Samuel Viggiano, David Oniani, Shyam Visweswaran, and Yanshan Wang. 2024. “Extraction of Sleep Information from Clinical Notes of Alzheimer’s Disease Patients Using Natural Language Processing.” Journal of the American Medical Informatics Association 31 (10): 2217–27. 10.1093/jamia/ocae177.

[6] Li, Z., Hu, Y., Lane, S., Selek, S., Shahani, L., Machado-Vieira, R., … Huang, M. (2024). Suicide Phenotyping from Clinical Notes in Safety-Net Psychiatric Hospital Using Multi-Label Classification with Pre-Trained Language Models. arXiv [Cs.CL]. Retrieved from http://arxiv.org/abs/2409.18878

[7] Team, G., Riviere, M., Pathak, S., Sessa, P. G., Hardin, C., Bhupatiraju, S., … Andreev, A. (2024). Gemma 2: Improving Open Language Models at a Practical Size. arXiv [Cs.CL]. Retrieved from http://arxiv.org/abs/2408.00118

[8] Pandit, S., Xu, J., Hong, J., Wang, Z., Chen, T., Xu, K., & Ding, Y. (2025). MedHallu: A Comprehensive Benchmark for Detecting Medical Hallucinations in Large Language Models. arXiv [Cs.CL]. Retrieved from http://arxiv.org/abs/2502.14302

[9] Sirrianni, J. W., Calloway, A., Hussain, S.-A., Chisolm, D., Kelleher, K., Seixas, A., Liu, H., Bartlett, C., & Davenport, M. A. (2025). Development of a Rule-Based Natural Language Processing Algorithm to Extract Sleep Information in Pediatric Primary Care Patients with a Sleep Diagnosis. medRxiv. 10.1101/2025.05.31.25328640

[10] Hussain, S.-A., Calloway, A., Sirrianni, J., Fosler-Lussier, E., & Davenport, M. (2025). Empirical Review of LLM-driven Classification of Multidimensional Sleep Health Mentions from Free-Text Clinical Notes. medRxiv. doi:10.1101/2025.06.04.25328983

